# Biting Diptera-host network structure varies with anthropogenic landscape modification

**DOI:** 10.64898/2026.05.05.26352205

**Authors:** Ben Bellekom, David R. Hemprich-Bennett, Naana Afua Acquaah, Bernard Adams, Ezekiel Donkor, Fred Aboagye-Antwi, Owen T. Lewis, Talya D. Hackett

## Abstract

1. Rapid and ongoing anthropogenic habitat modification has the potential to alter the species composition, abundance and activity of biting insect communities, which are important disease vectors. The resulting changes in the network of interactions between biting insects and their hosts have implications for the transmission of vector-borne pathogens.
2. We used DNA metabarcoding of Diptera blood meals to document bipartite networks of interactions between biting flies (Diptera) and their hosts (including humans, domesticated and wild animals) across a gradient of anthropogenic habitat modification (village, agricultural and near-natural habitat) surrounding two rural villages in Ghana.
3. We collected 7,095 biting Diptera (of 42 species) from 30 collection sites, and generated sequencing data from 75 blood meals (from 29 species). These blood meals contained DNA from 18 vertebrate host species, dominated by humans and their livestock.
4. Habitats with lower levels of anthropogenic modification had higher richness of biting Diptera and their host species. Species diversity and evenness did not differ significantly among habitats. Less modified habitats had higher network specificity, but connectance was highest in heavily modified habitats.
5. Humans were highly embedded within biting Diptera-host networks, detected in 68% of blood meals. The networks reveal several potential disease transmission pathways linking competent vectors with susceptible hosts. The presence of mixed blood meals containing DNA of both human and wild animal origin highlights the potential for transmission of established and emerging zoonotic disease via bridge vectors. The high betweenness-centrality within interaction networks of the important disease vector *Culex watti,* combined with its high abundance across all levels of anthropogenic landscape modification, suggest that it may be a connector species, linking and facilitating disease transmission between spatially distinct communities.
6. *Synthesis and applications:* Our results are of epidemiological interest, as they identify the exposure of humans to pathogen transmission cycles across a gradient of anthropogenic habitat modification through the movement of opportunistic bridge vectors. We discuss the implications for the transmission of emerging and established zoonotic disease and for the targeting and implementation of initiatives to reduce disease exposure and transmission.

## Introduction

Landscape modification characteristic of the Anthropocene can directly and indirectly drive changes in communities at the boundary between modified and natural habitats, increasing the risk of pathogen transmission. Biting Diptera, feeding as adults on the blood of vertebrates, transmit a wide range of pathogens that affect humans, as well as both wild and domestic animals, and represent a significant threat to public and economic wellbeing (Braack et al., 2018). Globally, an estimated one-sixth of all illness and disability is linked to vector-borne diseases, with over half of the global population currently at risk of infection (Campbell-Lendrum et al., 2015). Low- and Middle-income countries (LMIC) in the Global South are disproportionately at risk, with over 831 million people (70% of the population) at risk of infection from vector borne diseases in Africa alone (Agboli et al., 2021; Benelli and Beier, 2017). These risks are linked to extensive on-going changes in natural landscapes for agricultural and urban expansion (Chaves et al., 2020; Jones et al., 2013).

Anthropogenic landscape modification increases contact between humans, domestic and wild animals, and associated vectors, impacting epidemiological processes (Gibb et al., 2020b; Mayi et al., 2019; Patz et al., 2004). Rural populations that rely on subsistence farming are especially at risk, because vector abundances are positively associated with the primary sources of rural income (especially livestock) (Franklinos et al., 2019). Rural populations are also often situated adjacent to sylvatic habitats, allowing pathogens to cycle between wild animals and vectors, increasing exposure to established and emerging zoonoses (Gibb et al., 2020a). Anthropogenic land-use change fragments the structure of the landscape, producing smaller patches of sylvatic habitat surrounded by agricultural or urban land (Chaves et al., 2010). The resulting increase in edge habitats increases interaction frequencies and increases the potential for opportunistic biting Diptera species to act as bridge vectors (Hoyos et al., 2021; Meyer Steiger et al., 2016). Furthermore, host species diversity declines as human land-use intensifies (Abella-Medrano et al., 2018). This can increase disease transmission potential by eroding the dilution effect (where high host diversity limits the probability of a vector encountering a competent host (Civitello et al., 2015; Miller and Huppert, 2013)). Consequently, habitats with limited host diversity (e.g. urban or agricultural land) likely have higher zoonotic transmission and spillover, especially if these habitats are dominated by competent intermediate or ‘amplifier’ hosts, within which a pathogen may replicate rapidly and be maintained at a high concentration (Franklinos et al., 2019; Jones et al., 2013). Landscape modification can also alter the diversity and composition of biting Diptera communities by changing aspects of the biotic and abiotic environment, such as the availability of standing water, vegetation cover and host availability (Chaves et al., 2021; Ferraguti et al., 2016; Li et al., 2014). Depending on their particular lifecycles and habitat requirements, different biting Diptera species are likely to vary in their responses to such landscape changes (Perrin et al., 2022). For example, for species with aquatic larvae, increased local temperatures and artificial water bodies providing breeding habitats outside the normal seasonal rainfall periods in agricultural and village habitats decreases larval development time and allows reproduction independent of seasonal precipitation cycles (Gratz, 1999; Kesavaraju et al., 2008; Mattah et al., 2017). Human land-use is thus expected to alter and homogenise assemblages of biting Diptera and their vertebrate hosts, increasing transmission and spillover potential and altering networks of trophic interactions (Chaves et al., 2021; Ferraguti et al., 2016; Van Hoesel et al., 2019).

Despite the intimate and important interconnections among humans, livestock, biting Diptera and pathogens (Bellekom et al., 2021), empirical interaction data allowing for quantitative network analyses remains limited, particularly in West Africa. Here we investigate interactions of biting Diptera with their hosts in two rural villages in Ghana. Increasing anthropogenic landscape modification, expansion in agricultural land, and human encroachment into sylvatic habitats in recent decades (Food and Agriculture Organization of the United Nations, 2023; Acheampong et al., 2019) mean that a significant proportion of the Ghanaian population now live and work at the interface between rural and sylvatic habitat, with implications for the community structure of biting Diptera, their associated host interactions, and disease transmission potential.

We use PacBio Sequel sequencing of Diptera bloodmeals to investigate how networks linking biting Diptera (including known or potential disease vectors) to their vertebrate hosts differed between anthropogenically-modified habitats. We test the hypothesis that anthropogenic landscape modification would homogenise the community of vectors and hosts, altering the structure of bipartite interaction networks, and increasing the centrality of humans within them.

## Materials and Methods

### Study sites

We sampled insects around two rural villages within the Volta region, Ghana: Abutia Amageme (AA) (6.459064°N, 0.316247°E) and Mafi Agove (MA) (6.208310°N 0.442600°E; **Error! Reference source not found.**). Both are small, farming-dependent villages with cultivated land primarily used for maize and cassava surrounded by a semi-natural mixture of forest and grassland. Abutia Amegame retains more natural and semi-natural vegetation than Mafi Agove, which is larger with a less heterogeneous landscape matrix; see (Hemprich-Bennett et al., 2026) for full site descriptions. No biting Diptera control initiatives were in place during the sampling period, although some residents sleep under insecticide-treated bed nets.

### Insect collection

Insect sampling at each village took place between July 2019 and February 2020, over a 24-hour period every three weeks. On each visit, we sampled at four locations adjacent to each village. Each location was positioned randomly each week within each of four quadrants (NE, NW, SE, SW) of a 500 m radius circle centred on the village. We captured biting Diptera using CDC (Centre for Disease Control and Prevention) light traps (Bioquip, Rancho Dominguez, CA) 1.3 metres above the ground and modified to include a collection chamber filled with 95% ethanol (Bellekom et al., 2023; **Error! Reference source not found.**), with one trap at each of the four sampling locations. We used a sugar fermented yeast CO_2_ bait, consisting of 1L of water, 100g of sugar and 7g of yeast within a 1.5L bottle, attached to the trap entrance (Supplementary Fig. 2; Jerry et al., 2017; Smallegange et al., 2010).

Anthropogenic habitat modification at each sampling location was assessed using a combination of field-based assessment of vegetation type and Google Earth satellite imagery. Locations where cultivated land or livestock were the dominant land-use were categorised as Agricultural; those containing natural vegetation with limited human presence were classified as Near-natural; and sampling locations in or around human habitation were classified as Village. Data were accumulated from 30 collection sites: 12 Agricultural, 12 Near-natural, and six Village.

### Sample processing

Insect samples were stored soon after collection and for up to 48-hours in a refrigerator and then transported in cold boxes to a freezer at the University of Ghana, Accra. DNA amplification success from bloodmeals stored in 95% ethanol is not strongly affected by storage temperature (Bellekom et al., 2023), however, time at room temperature was still kept to a minimum. Prior to molecular identification, samples were visually sorted to Order and assigned a morphospecies. Up to five representatives of each morphospecies were placed in 96-well microplates containing 95% ethanol, and the total abundance of each morphospecies per sampling event was recorded. Blood-fed individuals (those with a visible blood meal) and potentially blood-fed individuals (gravid and parous individuals, and those with almost entirely digested blood meals) were visually identified and separated into a different 96-well plate for blood meal analysis. All plated samples were shipped under permit in cold boxes to the Centre for Biodiversity Genomics facility at the University of Guelph for DNA sequencing.

### Blood meal analysis

Samples were processed using automated pipelines established by the Canadian Centre for DNA Barcoding. Diptera and blood meal DNA were handled by a Biomek FXP liquid handling system and extracted using a glass fibre extraction protocol (Ivanova et al., 2006). Single molecule real-time (SMRT) sequencing was carried out on the PacBio Sequel II platform, targeting the COI gene. Amplicons were generated using the SMRTbell Express Template kit 2.0, Sequel II Binding kit 2.1 and Sequel II DNA Internal Control 1.0 (Hebert et al., 2018).

### Bioinformatics

Sequencing data were initially processed using the mBRAVE platform. Sequences were quality filtered and clustered into OTUs using default parameters, with a 2% sequence divergence threshold. Low-abundance OTUs (<2% of reads per sample) were excluded. Chimeric sequences, false reads, and controls were manually removed. A custom CO1 reference library was created using NSDPY (Hebert and Meglécz, 2022) and blastn (Wheeler and Bhagwat, 2007), incorporating families expected in the region based on a manual search of GBIF (gbif.org, 2021) and field guides (Grubb, 1998); (Borrow and Demey, 2010). Taxonomic assignment used blastn (word size = 28, max_target_seqs = 1) and a 98% sequence identity threshold.

As individual samples may generate multiple contigs, singleton contigs within mixed samples were removed as likely artefacts. Any singletons observed only once in the entire dataset were retained, as they were deemed to likely be true results rather than contamination. Where a sample was represented by multiple contigs with differing species assignments, contigs were retained only if they were not singletons of expected fragment length (∼180bp) and ≥98% identity match (Hopken et al., 2021). Where a sample could not be identified to species level, the highest taxonomic resolution was assigned. Samples that retained contigs of different species origin after filtering were classified as having a mixed blood meal. To minimise potential human contamination, contigs identified as human DNA were only retained if they contained >10 reads in a sample, based on observed read distributions of confirmed non-human host taxa (e.g. *Philantomba maxwellii*).

Blood-fed and non-blood-fed Diptera were processed using the same mBRAVE protocol. Where taxonomy could not be assigned automatically using mBRAVE, sequences were manually identified using BLAST. Where a sample could not be identified to species level, a unique identifier was assigned to prevent multiple species being represented by a single family identification in downstream analysis. Abundances were assigned to each occurrence of a species using the estimated abundance for each morphospecies. Where a morphospecies was represented by multiple species, the abundance of each component species was assigned based on their overall relative proportion.

### Data analysis

We explored the effect of habitat type on log-transformed Diptera Shannon-Wiener diversity using a generalized linear model with a Gaussian error distribution. The statistical significance of habitat type was assessed by comparing nested models for variation in deviance based on a chi-square distribution (Mayi et al., 2020). Post hoc analysis was conducted to identify differences between individual habitat types using a Tukey’s HSD (honestly significant difference) test. The dissimilarity of the biting Diptera community composition across habitat types and locations was investigated using a Permutational Multivariate Analysis of Variance Using Distance Matrices (PERMANOVA) (Oksanen et al., 2025).

Interaction data were pooled across sampling locations by habitat type to generate a weighted bipartite network for each category. To examine the complexity of our networks, we calculated their connectance, the fraction of potential ecological interactions that are realised (Poisot and Gravel, 2014). Network specialisation for each habitat classification was examined using H2’, which quantifies the deviation of observed interaction frequencies from the frequencies expected if interactions are random (Blüthgen et al., 2006). As generalist species are more likely to act as ‘connector species’ (Hackett et al., 2019) that promote cross-species disease transmission, networks with a high degree of generality may sustain a greater disease transmission potential between unrelated hosts (Su et al., 2022). To assess the risk of disease exposure potential for humans in each habitat type, we calculated the degree and closeness centrality for humans in each network. As the within-species prevalence of a disease increases linearly with its degree, nodes with a high degree (i.e., many connections to the target node) are more likely to participate in disease transmission events (Su et al., 2022). Closeness centrality describes the mean path length from the target node to all other nodes in the network, with a high value indicating that the node may be rapidly affected by other nodes in the network and vice versa (Silk et al., 2017; Su et al., 2022). To examine the relative importance of a node in connecting parts of each network and their potential to mediate transmission throughout the network, we calculated the betweenness centrality of the biting Diptera in each of our networks. Betweenness centrality describes the proportion of shortest pathlengths in a network that go via the node, with a high value indicating that the node is important for the cohesiveness of the network (Llaberia-Robledillo et al., 2022). While we did not directly identify pathogens within the blood meals, we inferred disease vectoring potential of each blood-fed Diptera species using Vectorbase (GiraldoCalderón et al., 2015) to highlight potential transmission events for susceptible hosts. To assess sampling completeness, we constructed interpolation and extrapolation curves for hosts, biting Diptera and their interactions as a function of sampling effort (the number of blood meals collected) using the iNEXT package (q=0, data type = incidence frequency) (Hsieh et al., 2016). Finally, we estimated total interactions, and host and Diptera species richness using the ChaoRichness function (Hsieh et al., 2016), which is based on methods proposed by (Chao, 1987).

## Results

### Overview

In total we collected 7,095 biting Diptera from the 30 collection sites (12 Agricultural, 12 Near-natural, and six Village sites) (**Error! Reference source not found.**, **Error! Reference source not found.**). We identified 42 individual biting Diptera species (84% of nodes) and resolved an additional eight biting Diptera to genus or family level (16% of nodes) (**Error! Reference source not found.**1). Blood-fed individuals were collected from 20 sampling locations: 6 Agricultural, 11 Near-natural, and 3 Village. Blood meal sequencing on the PacBio Sequel II platform generated an initial 189,240 reads. After filtering and removal of chimeras this was reduced to 184,938 reads. Approximately half (95,772) of our reads were assigned to a BIN, whilst 89,166 were placed in 11,016 OTUs. Following progression through our BLAST pipeline, using our custom reference library for host species identification, this was reduced to 111,449 reads. After implementing our strict filtering criteria, we retained 3,151 reads which characterised the composition of 75 blood meals and identified 18 individual host species derived from blood meals of 29 biting Diptera species.

### Effect of habitat modification

Non-blood-fed and blood-fed Diptera richness and abundance was lower in Abutia Amegame than in Mafi Agove (**Error! Reference source not found.**2). Total richness and abundance of non-blood-fed and blood-fed Diptera were lower in the Village habitats compared to Agricultural and Near-natural habitat, as was host richness (Table 1: The proportion of fed individuals for each biting Diptera species sampled across all sites.

**Table 1:** The proportion of fed individuals for each biting Diptera species sampled across all sites.

| Species | Total | Blood-fed | Proportion blood-fed |
| --- | --- | --- | --- |
| *Culex nebulosus* | 4 | 1 | 25.00% |
| *Culex watti* | 175 | 24 | 13.71% |
| *Aedes bromeliae* | 2 | 1 | 50.00% |
| *Culex theileri* | 2 | 0 | 0.00% |
| *Aedeomyia africana* | 2 | 0 | 0.00% |
| *Culicoides imicola* | 1649 | 2 | 0.12% |
| *Culicoides hortensis* | 130 | 3 | 2.31% |
| *Sergentomyia ingrami* | 17 | 1 | 5.88% |
| *Sergentomyia hamoni* | 18 | 1 | 5.56% |
| *Sergentomyia africana* | 227 | 1 | 0.44% |
| *Culex rima* | 9 | 3 | 33.33% |
| *Mimomyia mimomyiformis* | 12 | 7 | 58.33% |
| *Coquillettidia metallica* | 20 | 2 | 10.00% |
| *Culicoides distinctipennis* | 2766 | 2 | 0.07% |
| *Mimomyia mediolineata* | 4 | 0 | 0.00% |
| *Culex perexiguus* | 8 | 2 | 25.00% |
| *Sergentomyia inermis* | 337 | 1 | 0.30% |
| *Culex aurantapex* | 2 | 0 | 0.00% |
| *Mansonia africana* | 25 | 8 | 32.00% |
| *Culex decens* | 2 | 1 | 50.00% |
| *Forcipomyia sp. 1* | 347 | 0 | 0.00% |
| *Culicoides oxystoma* | 130 | 0 | 0.00% |
| *Sergentomyia schwetzi* | 40 | 1 | 2.50% |
| *Sergentomyia sp. 1* | 41 | 1 | 2.44% |
| *Dasyhelea ludingensis* | 555 | 0 | 0.00% |
| *Sergentomyia clydei* | 15 | 0 | 0.00% |
| *Culex univittatus* | 7 | 1 | 14.29% |
| *Psychoda sp. 1* | 140 | 0 | 0.00% |
| *Culex tritaeniorhynchus* | 7 | 1 | 14.29% |
| *Culex cinereus* | 1 | 1 | 100.00% |
| *Aedes unilineatus* | 3 | 0 | 0.00% |
| *Uranotaenia alboabdominalis* | 3 | 0 | 0.00% |
| *Aedes hirsutus* | 4 | 0 | 0.00% |
| *Culex duttoni* | 2 | 0 | 0.00% |
| *Culex naevei* | 3 | 0 | 0.00% |
| *Anopheles coustani* | 4 | 0 | 0.00% |
| *Culex vansomerini* | 4 | 1 | 25.00% |
| *Culex adersianus* | 2 | 0 | 0.00% |
| *Aedes quasiunivittatus* | 2 | 0 | 0.00% |
| *Culicoides neavei* | 3 | 0 | 0.00% |
| *Uranotaenia bilineata* | 2 | 0 | 0.00% |
| *Culex annulioris* | 2 | 0 | 0.00% |
| *Culicoides enderleini* | 257 | 0 | 0.00% |
| *Ceratopogonidae sp.1* | 4 | 1 | 25.00% |
| *Sergentomyia distincta* | 12 | 3 | 25.00% |
| *Ceratopogonidae sp.2* | 45 | 1 | 2.22% |
| *Ceratopogonidae sp.3* | 42 | 1 | 2.38% |
| *Uranotaenia micromelas* | 3 | 1 | 33.33% |
| *Atrichopogon sp.1* | 4 | 1 | 25.00% |
| *Allohelea sp.1* | 1 | 1 | 100.00% |

**Table 1:** Non-blood-fed and blood-fed Diptera richness, number of blood meals, and host richness by level of anthropogenic landscape modification.

| Site/Habitat | Non-blood-fed Diptera richness | Non-blood-fed Diptera abundance | Blood-fed Diptera richness | Number of blood meals | Host richness |
| --- | --- | --- | --- | --- | --- |
| Village | 11 | 1,382 | 4 | 8 | 2 |
| Agricultural | 29 | 1,454 | 11 | 28 | 9 |
| Near-natural | 34 | 4,259 | 22 | 39 | 11 |

Table 1). Biting Diptera species evenness was similar (F_2,20_ = 1.570; p = 0.233) in Village (mean = 0.109, SE = 0.04), Agricultural (mean = 0.272, SE = 0.06) and Near-natural (mean = 0.220, SE 0.04) habitats. There was no significant effect of habitat modification on biting Diptera diversity (χ^2^ = 0.29178; p = 0.186) (Figure 1).

**Figure 1:**
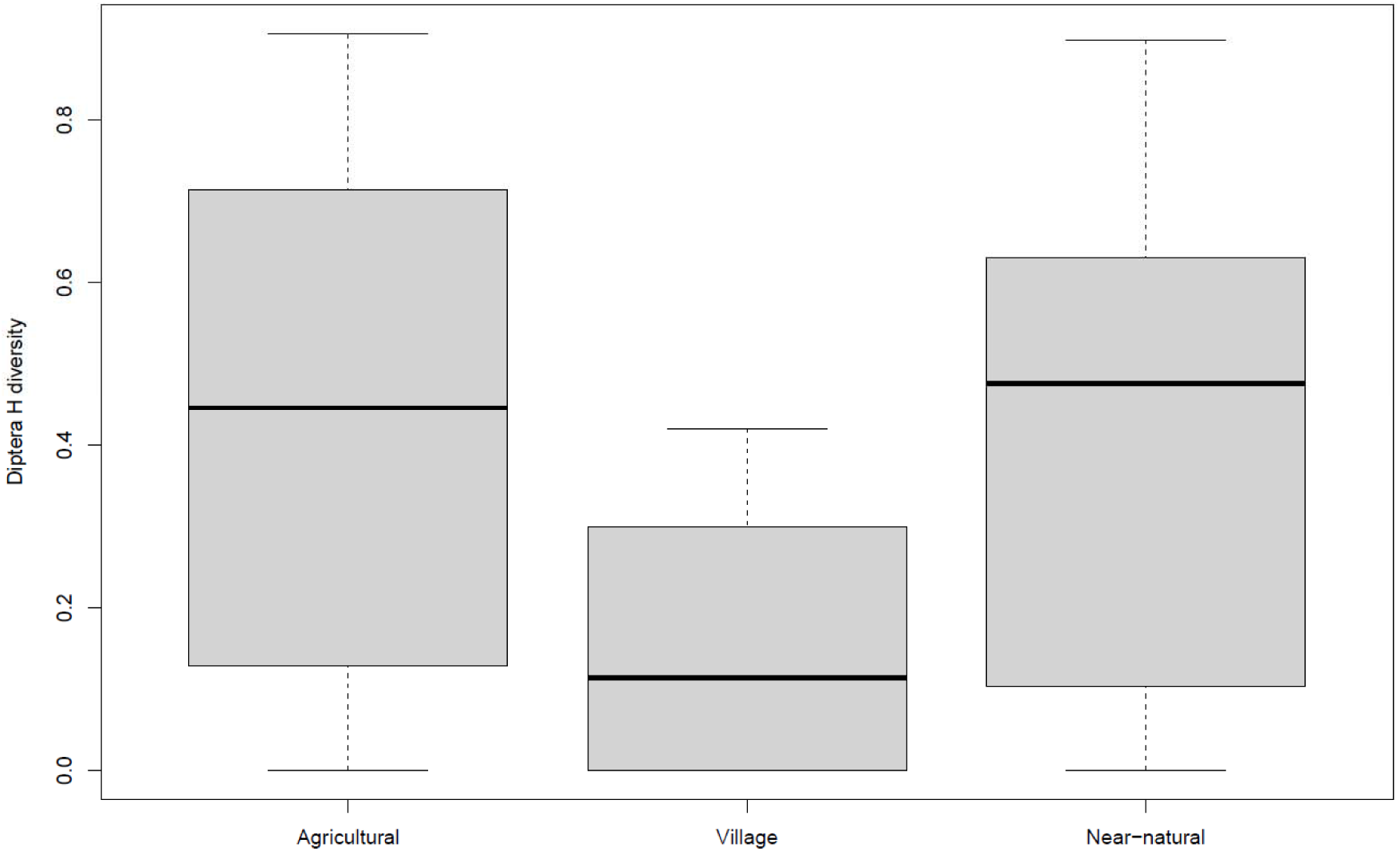
Biting Diptera Shannon (H) diversity by habitat type. Each box displays the interquartile range, and the solid line represents the median. Whiskers display the maximum and minimum interaction evenness and H values for each habitat type

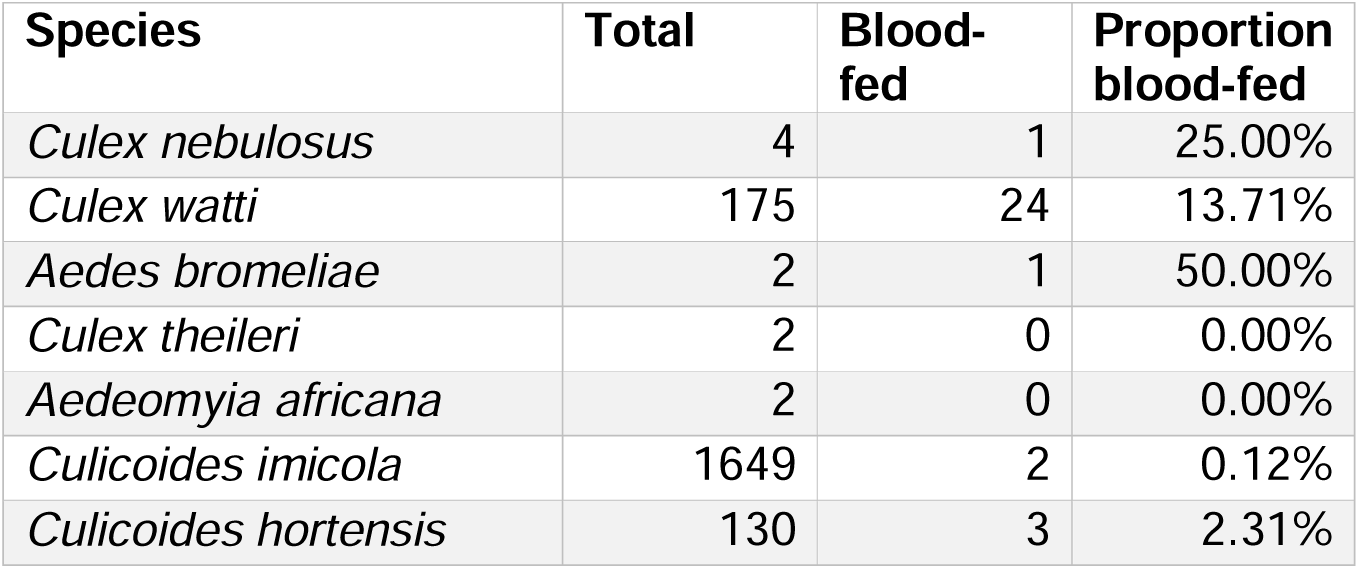
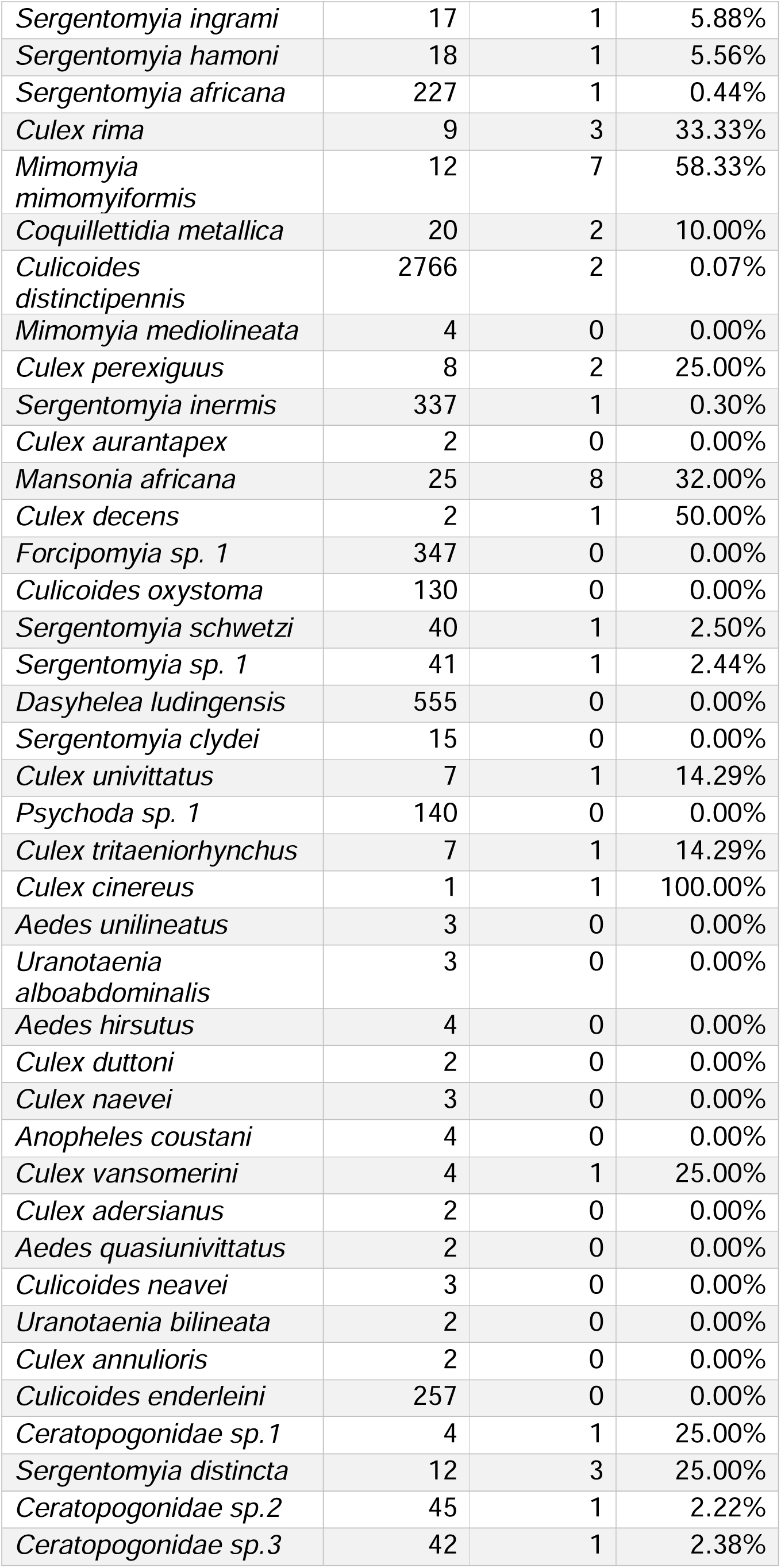
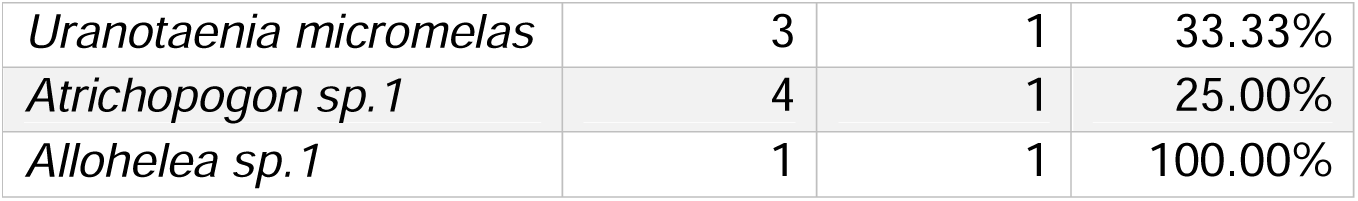

Biting Diptera community composition did not differ significantly across habitat types (F_2,27_ = 1.09; p = 0.306; Figure 2A) or site (F_1,28_ = 0.862; p = 0.611; **Error! Reference source not found.**).

**Figure 2:**
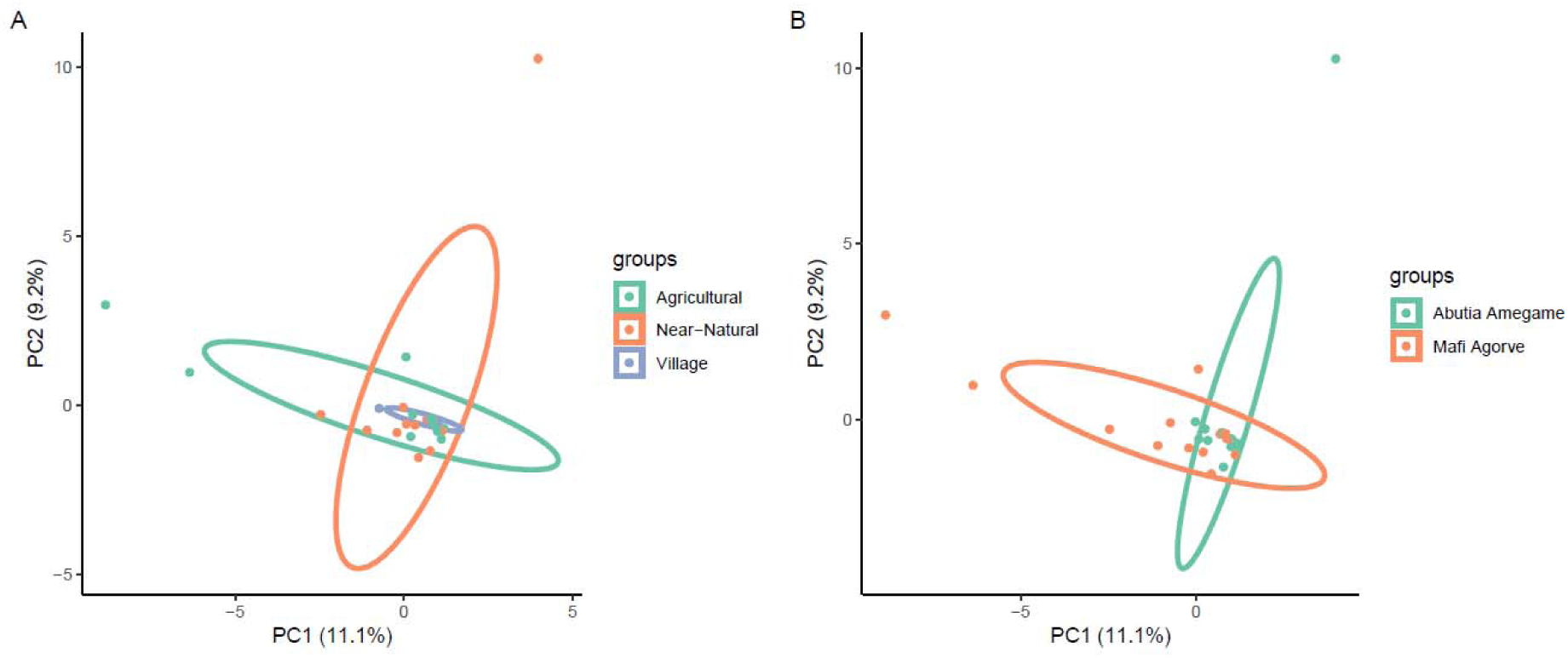
Ordination of the overall biting Diptera community composition by A) habitat category or B) Village. Each point represents the biting Diptera diversity captured by a unique sampling event in Village (purple) Agricultural (blue), and Near-natural (orange) habitat. Or in Abutia Amegame (blue) and Mafi Agove (orange).

### Blood meal origin

Humans had the highest number of interactions across all sites and habitat classifications; human DNA was detected in 68% of all collected blood meals. The number of wild host species differed markedly between the villages, with 10 species in Mafi Agove and just one in Abutia Amegame, with minimal difference in domestic animal species richness (3 and 4 species, respectively). There were slightly more domestic and wild host species in Near-natural habitat (4 and 6) than in Agricultural habitat (3 and 5). In Village habitats we only identified interactions with human, and one mixed blood meal containing human DNA and DNA from a wild bird species (a quail, *Coturnix* sp.).

Mixed blood meals accounted for 7.89% (6) of all blood meals. Three mixed blood meals were found in samples from Near-natural habitat (7.69%), two from Agricultural habitat (7.14%) and one from Village (12.5%). Five of the six mixed blood meals contained human DNA. Two of the mixed blood meals contained DNA from three host origins (Soricidae, Fringillidae, and Phasianinae) and (*Capra hircus*, *Ovis aries*, and *Homo sapiens*).

### Potential disease transmission events

Of the 29 blood-fed biting Diptera species collected, 18 (62%) were competent of vectoring one or more pathogen of medical or veterinary importance. Of the vector-competent Diptera species, 16 (88%) had interacted with at least one host species susceptible to the pathogens they vector (Figure 3; Supplementary Table 3).

**Figure 3:**
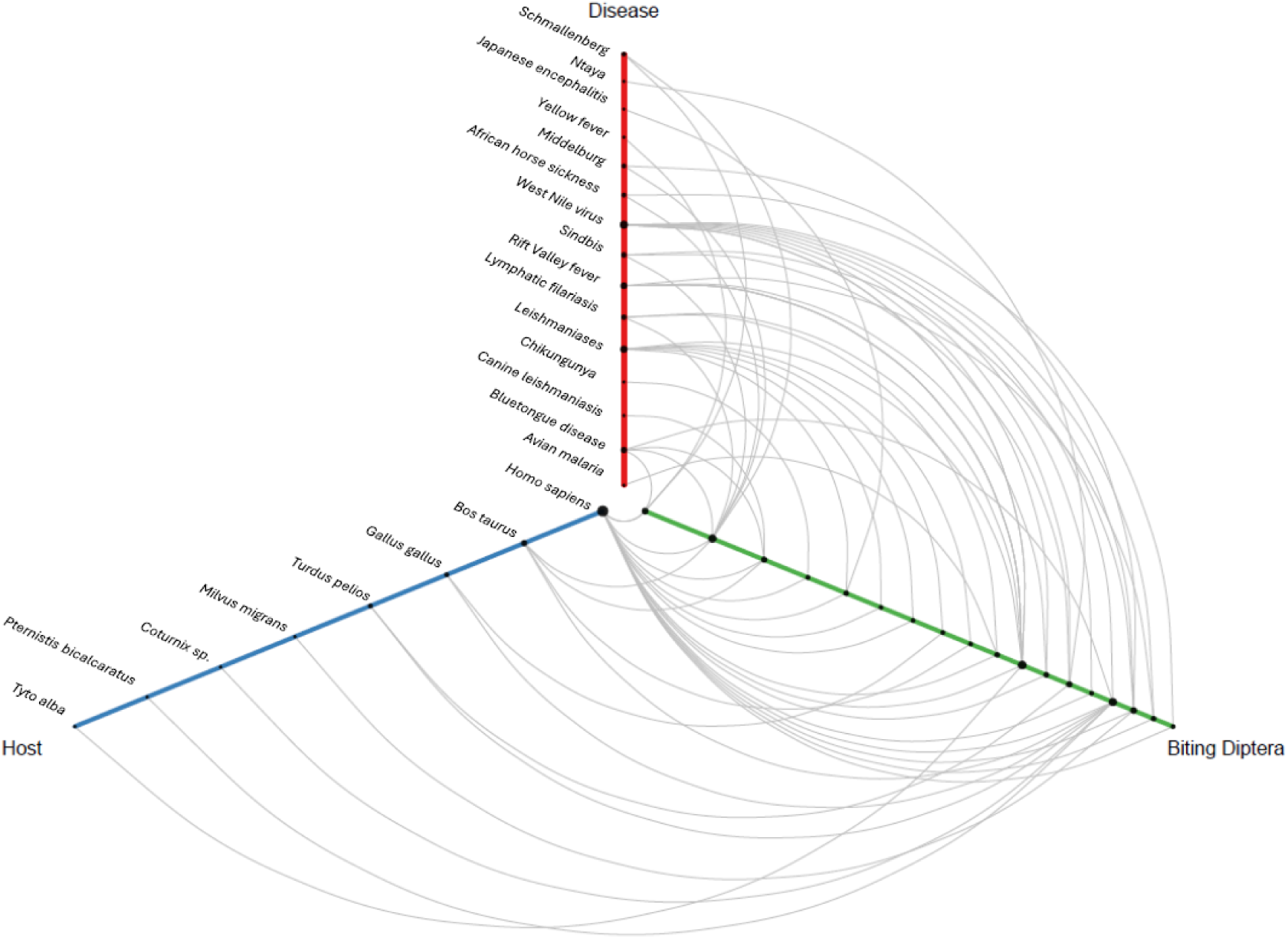
Hive plot showing how potential transmission events between hosts (blue), biting Diptera (green) and their respective diseases (red). Node size along each axis scale with frequency. Diptera nodes, from inside to outside, are: *Culex perexiguus, Culicoides imicola, Culicoides distinctipennis, Sergentomyia schwetzi, Aedes bromeliae, Sergentomyia ingrami, Sergentomyia Africana, Sergentomyia hamoni, Sergentomyia inermis, Sergentomyia distincta, Mansonia Africana, Culex decens, Culex univittatus, Culex cinereus, Culex watti, Coquillettidia metallica, Culex nebulosus, Culex vansomereni*.

### Community structure

Accumulation curves showed that host and biting Diptera species richness for Agricultural and host richness for Near-natural habitat were relatively well resolved, with curves approaching an asymptote in each case. Sampling for biting Diptera species in Near-natural habitat and species interactions in all habitats were incomplete (Figure 44 and **Error! Reference source not found.**). Due to the lack of interaction data, Village habitats were excluded from sampling completeness analysis.

All interaction data were compiled into an overall summary network (Figure A), and separate networks were also constructed for each habitat category (Fig. 5B-D) and each location (Supplementary Fig. 4). The connectance of the overall network was 0.088. Whilst no statistical comparison of network metrics across habitats was possible due to lack of replication, the Village network had higher connectance (0.625) than either the Agricultural (0.191) or the Near-natural (0.123) network, although sample sizes in terms of numbers of blood meals were very low for the Village sites.

**Figure 4:**
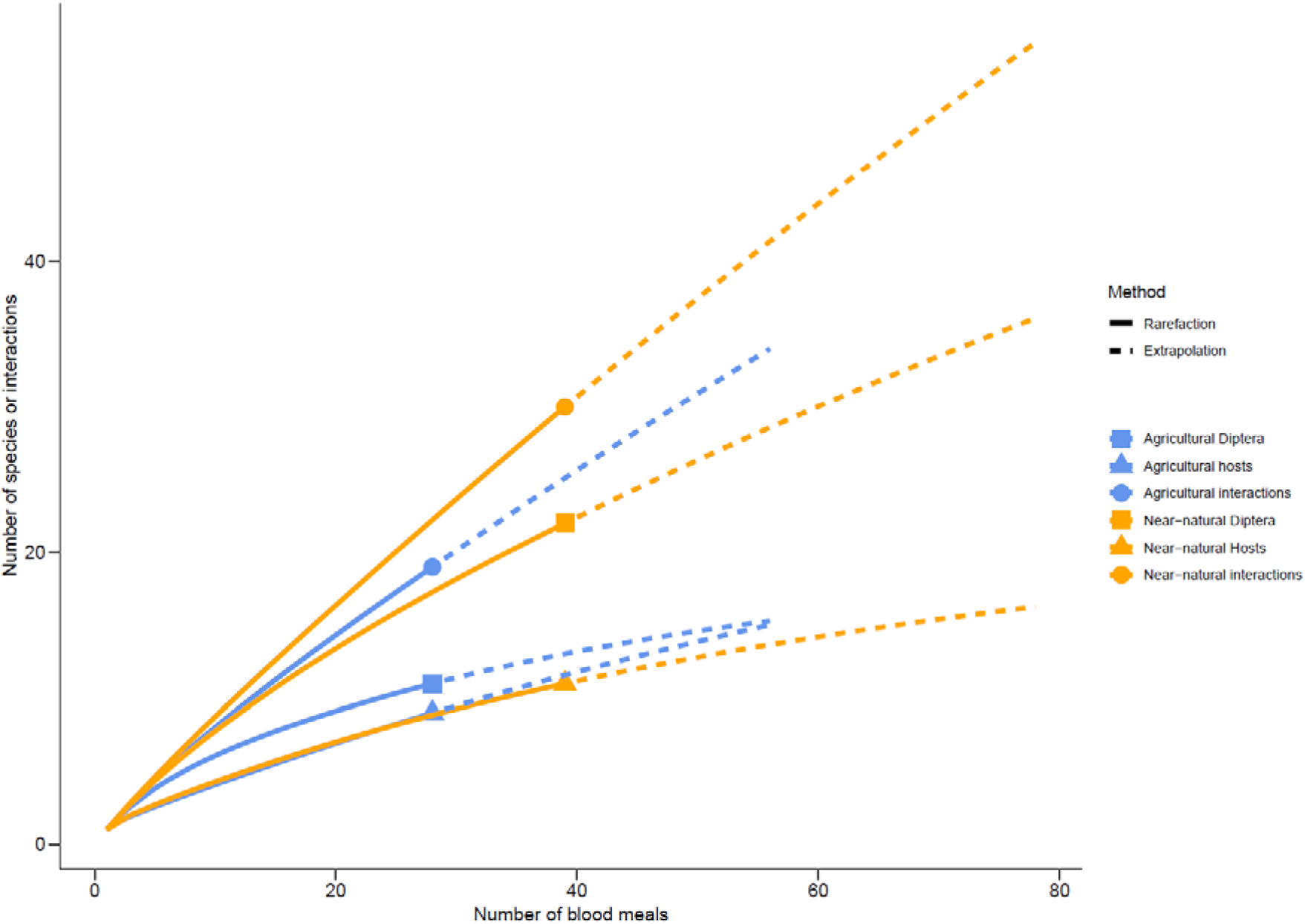
Smoothed accumulation and extrapolation curves to assess sampling completeness. Total numbers of hosts (triangle), biting Diptera (square), and interactions recorded in the whole dataset (circle), by habitat type (Agricultural in blue, Near-natural in orange), as a function of sampling effort (the number of blood meals screened). Village habitat was excluded from this analysis due to under sampling

**Figure 5:**
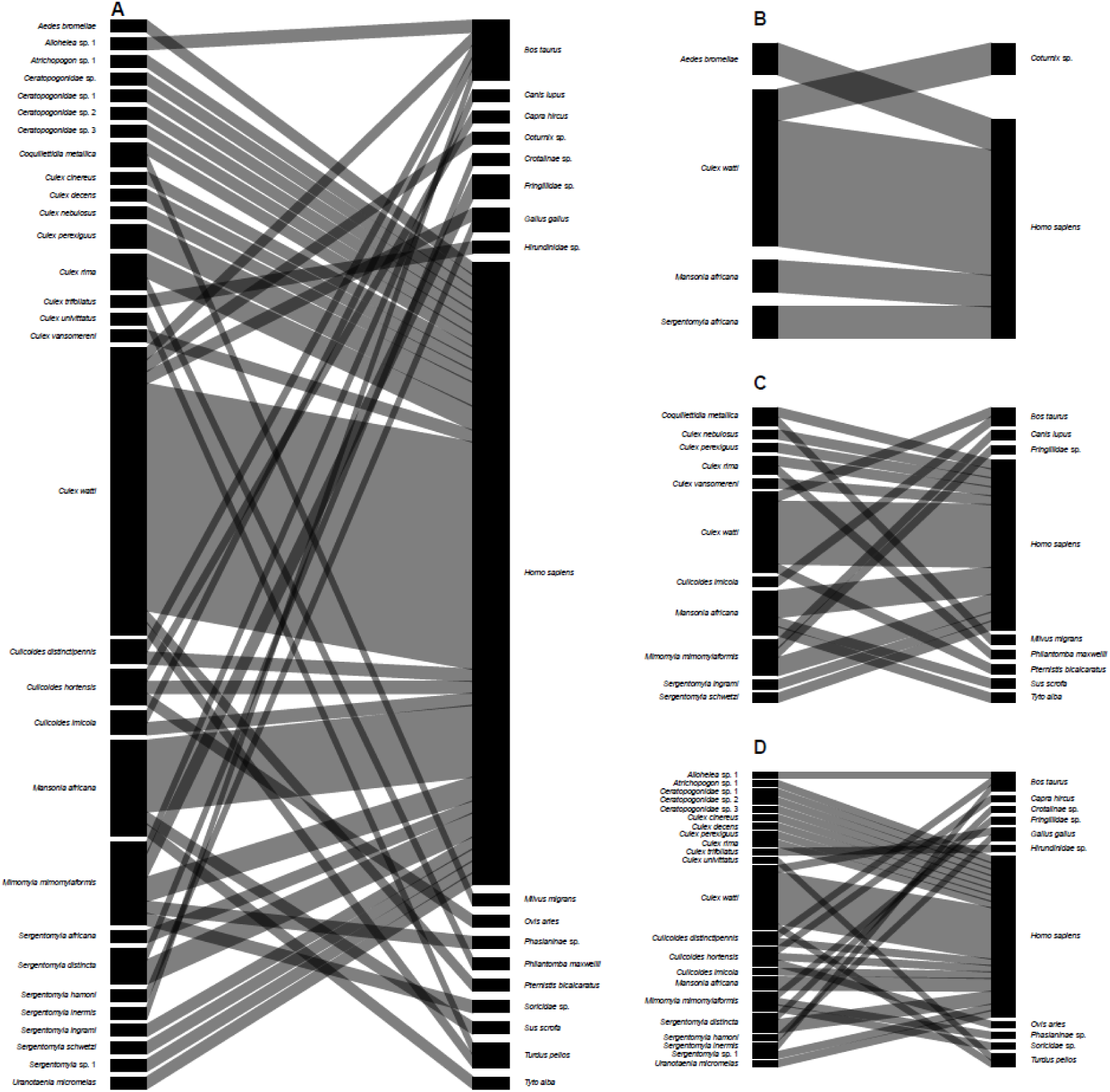
Combined habitat networks. (A) The aggregated network containing all host (right) and Diptera (left) interactions, interactions separated by habitat classification: Village (B), Agricultural (C), and Near-natural (D). Node and edge widths are proportional to frequency of occurrence

The H2’ specialisation of the combined network across both sites was 0.192, and network specialisation was higher in Near-natural (0.347) than Agricultural (0.148) habitats. There were insufficient data to reliably calculate the Village H2’ value.

Human closeness centrality was 0.094 and degree 25. Closeness centrality was higher in Village habitat (0.5) than Agricultural (0.170) and Near-natural habitat (0.171). The degree of humans was higher in Near-natural habitat (16) than in Agricultural (10) and Village habitats (4).

*Culex watti* had the highest betweenness centrality of all the blood-fed biting Diptera in our combined network (0.450). Moreover, *C. watti* had the highest betweenness centrality across both Near-natural (0.507) and Agricultural (1) habitats. Due to the lack of data, Village was excluded from this analysis.

## Discussion

Our results revealed variations in the richness of biting Diptera across a gradient of anthropogenic habitat modification, with higher richness and abundance in Near-natural and Agricultural habitats. Network metrics also differed with anthropogenic landscape modification, and the detailed data on individual vector species and their feeding interactions that emerge from our networks reveal a wide variety of potential transmission pathways between biting Diptera and disease-competent hosts.

Ecological networks populated with interaction data generated through the DNA sequencing of biting Diptera blood meals provide an insight into the community and interaction structure of Diptera and their hosts in a sampled location. Whilst every effort was taken to reduce bias, there are several potential limitations to this approach. Interaction networks provide a snapshot of community and interaction structure, and multiple years of sampling are required to document temporal changes in network structure (Olivier et al., 2019). To minimise the effect of varying diel activity patterns, for each sampling event we sampled over a period of 24 hours. Under-sampling may alter conclusions drawn from several network metrics (Blüthgen, 2010); the rarefaction analysis demonstrated that interactions, and host and Diptera richness, are indeed under-sampled. Significantly increased sampling effort, beyond what was possible within time and travel limitations, would be required to increase the confidence in the network metrics values (Ings et al., 2009)

Overall abundance of biting Diptera decreased with landscape modification, with a biting Diptera abundance nearly three times higher in Near-natural habitat compared to other habitats. This could reflect the proximity and availability of breeding sites and hosts (Barrientos-Roldán et al., 2022; Ferraguti et al., 2016; Johnston et al., 2014; Pereira-Silva et al., 2021; Young et al., 2021). Indeed, despite highly modified land, such as Village habitat, providing breeding opportunities for biting Diptera outside of seasonal precipitation cycles, only those synanthropic species capable of propagating and feeding on hosts in these habitats are likely to be present (Barata et al., 2012; Johnston et al., 2014). Consequently, the higher biting Diptera species richness and abundance in Near-natural and Agricultural habitat are likely due to the habitat providing a greater number of hosts and more variable breeding sites that support more species and higher abundances of each species. The observed pattern of increased host richness within blood meals in Near-natural habitat is consistent with the expectation that Near-natural habitats retain more wild species. This host diversity could help reduce disease transmission in less modified habitat through the dilution effect (Shah et al., 2019).

Species evenness was low across all habitats, but was lowest in village interiors, consistent with previous studies (Möhlmann et al., 2017; Petruff et al., 2020; Visa Shalini et al., 2022). Several species were abundant across all habitat types, including *Culicoides imicola*, *Culicoides distinctipennis*, *Sergentomyia africana* and *Culex watti*. These species are likely habitat generalists with a degree of plasticity in blood feeding behaviour, allowing them to exploit a wide range of hosts (Stone and Gross, 2018; Villard and Metzger, 2014). Indeed, we recorded *Culex watti* as having blood meals from six host species. As these abundant species are competent vectors of a range of pathogens, their abundance and wide distribution have significant implications for the emergence of zoonotic diseases and disease transmission potential across a range of habitats.

We found no significant dissimilarity in community composition across habitats. However, we identified biting Diptera species that were unique to each habitat. Indeed, 53% of Diptera species collected in the Near-natural classification were unique to this habitat, perhaps reflecting a restriction to habitat conditions absent in human-modified parts of the landscape (Vittor et al., 2009). Communities in transitional zones (ecotones) often comprise species from the adjacent habitats (Fortin et al., 2000). We observed a pattern of high Diptera species overlap across an anthropogenic landscape modification gradient from Agricultural to Near-natural (34% of Agricultural species were also present in Near-natural habitat), indicating a shared community and consistent with movement between the two habitat types (Meyer Steiger et al., 2016). Overlapping biting Diptera communities link Agricultural with adjacent Near-natural habitat and, consequently, humans with the sylvatic cycle through the movement of opportunistic bridge vectors (Miot et al., 2020). We found several species that link the Village and Agricultural habitat, including *Anopheles coustani*. This species is a known vector of diseases including malaria, and displays anthropophilic behaviour (Ciubotariu et al., 2020). These findings are epidemiologically relevant as they indicate that existing and future Agricultural habitat may facilitate disease transmission across the landscape and provide a pathway through which emerging and established zoonotic disease are introduced into rural and urban transmission cycles.

Network connectance was greater within the Village habitat than Agricultural and Near-natural habitats, although this may reflect the limited number of recorded interactions and interaction partners, and the abundance of interactions with humans (Valdovinos et al., 2009). Low network connectance in Agricultural and Near-natural habitat is likely a result of the increased host species richness (Rivera-Hutinel et al., 2012). Interpolation and extrapolation curves demonstrated that biting Diptera-host interactions were highly under-sampled in comparison to richness estimates, in all habitat types. Consequently, connectance values in our study may not be representative of the true interaction structure (Blüthgen, 2010; Heleno et al., 2012). Whilst most networks are incomplete (Vizentin□Bugoni et al., 2016), incomplete sampling, as in the current study, can still be sufficient to capture the majority of functionally important species in a network (Hegland et al., 2010).

Network specialisation (H2’) decreased with anthropogenic landscape modification, although values were uniformly low across habitats, likely reflecting plasticity in host use. Host usage has a high degree of plasticity in biting Diptera and may be strongly influenced by local host densities (Takken and Verhulst, 2013). Increased H2’ in Near-natural habitat could reflect the greater host and Diptera density, species richness, as well as potential niche partitioning (Chakravarty et al., 2023; Dalsgaard et al., 2011). Lower network specialism in Agricultural than Near-natural habitat is particularly epidemiologically relevant, as generalist vectors have an increased capacity to transmit emerging and established zoonoses into novel hosts (Santiago-Alarcon et al., 2012). Therefore, habitat with a lower network specialism will likely have greater disease transmission potential (Ellwanger and Chies, 2021). Indeed, agricultural drivers have previously been linked to over 50% of all zoonotic disease emergence (Rohr et al., 2019). Whilst each habitat network was considered distinct and sampling conducted over an extended period provided a snapshot of community structure in each habitat classification, the contiguous nature of the landscape, and overlapping Diptera communities, suggest that movement of individuals between habitats is likely. Such movement has the potential to link susceptible hosts in one network with competent vectors in another, facilitating disease transmission across the landscape.

The high degree centrality of humans in Agricultural and Near-natural habitat demonstrates that humans may act as a hub species (Toju et al., 2018), interacting with a wide range of biting Diptera species (including disease-competent vectors) and (indirectly) other hosts. Indeed, the presence of mixed blood meals containing DNA of human and wild animal origin highlights the potential for transmission of established and emerging zoonotic disease via bridge vectors (Brackney et al., 2021; Miot et al., 2020).

The closeness centrality of humans was greatest in Village habitat and there was minimal difference between Agricultural and Near-natural habitat, suggesting that transmission potential to humans is higher in anthropogenically modified habitat (Grubb et al., 2021; Ribeiro et al., 2020). These findings are particularly epidemiologically relevant, as they suggest that, at least in the study landscape, humans may be equally at risk of contracting vector-borne diseases when working in Agricultural land as they are when encroaching or traversing Near-natural habitat. The high centrality of humans in Village habitats is expected due to the anthropophilic feeding behaviour of the sampled blood-fed Diptera and their historical association with anthropogenic habitat (Bennett et al., 2015; Ughasi et al., 2012). However, high centrality may also reflect the limited number of blood meals collected from Village locations (Costenbader and Valente, 2003). Indeed, given the observed presence of domestic hosts, such as goats, chickens, and dogs, there was a surprising lack of domestic animal host DNA in the Village blood meals. This could in part reflect anthropophilic feeding behaviour, but is perhaps more likely a consequence of under-sampling of areas with high densities of dwellings, which were restricted to the centre of the two sampling areas. Despite lower closeness centrality values in Agricultural and Near-natural habitat, high human degree centrality in these habitats remains an epidemiological concern (Bell et al., 1999).

Across the range of blood-fed biting Diptera collected in all habitat classifications, *Culex watti*, a potential vector of West Nile Virus (Diarra et al., 2019), consistently had the highest betweenness centrality. A high betweenness centrality value indicates that the species is important for the cohesion of the network and favours the circulation of their vectored pathogens throughout the network (Espinaze et al., 2018; Llaberia-Robledillo et al., 2022). Whilst a higher number of blood-fed and non-blood-fed *C. watti* were collected in Agricultural and Near-natural habitats, the presence of *C. watti* within all habitat classifications indicates a degree of generality in their habitat range (Stone and Gross, 2018). Moreover, the high betweenness values and presence across all levels of anthropogenic landscape modifications suggest that *C. watti* has the potential to act as a connector species, linking and facilitating disease transmission between spatially distinct communities (Bellekom et al., 2021; Llaberia-Robledillo et al., 2022).

Increasing anthropogenic landscape modification could drive a range of vector-borne disease outbreaks and zoonotic emergence events through its effect on biting Diptera and host community composition and vector-host relationships (Meyer Steiger et al., 2016; Patz et al., 2004). The characterisation of the biting Diptera-host community and interaction structure provides opportunities to highlight and implement control initiatives to mitigate transmission potential. In the short term, such opportunities may include behavioural modification for village inhabitants to limit exposure to transmission, including informing locals of potential risk factors, limiting encroachment into Near-natural habitat, covering the skin and using insect repellent when working at the interface between Agricultural and Near-natural habitat to minimise biting, and increase uptake of insecticide-treated bed netting. Further, in the medium term, such interaction data may highlight vectors and hosts of particular importance to network cohesion and disease circulation. These data will inform localised control initiatives, for example, targeting synanthropic breeding habitat to suppress anthropophilic species, and the identification and removal of key domestic host species that may act as reservoirs of disease. In the long term, Diptera blood meal networks may be used to inform potential ecological consequences and network rewiring that may occur following vector control initiatives.

The high degree of overlap in community composition and shared host usage between Agricultural and Near-natural land places rural populations that rely on subsistence farming at the interface of disease transmission between rural and sylvatic habitat (Meyer Steiger et al., 2016). Closeness and degree centrality metrics indicated that there was a high disease transmission risk to humans. Incorporation of complementary quantitative susceptibility and vector competency rates may provide a more resolved assessment of vector-human transmission potential. Our interaction data, when combined with vector competency data, indicated a high number of potential transmission pathways. Subsequent interaction and pathogen data would thus allow for network approaches to be applied on an individual basis, in which nodes represent host or Diptera individuals and edges potential transmission events (Bellekom et al., 2021). Such data would be highly epidemiologically relevant and allow for the monitoring of the transmission of established and emerging zoonosis across the rural-sylvatic interface.

## Data Availability

Data is archived on the Zenodo digital repository: https://doi.org/10.5281/zenodo.19591599

## Acknowledgements

This research was supported by members of the Target Malaria Ghana Stakeholder Engagement Team, especially Divine Dzokoto and Linda Mawutor Aboagye. Data collection was facilitated by village elders, members of the two study communities, and especially those who assisted with trap deployment and sample collections. Sample sorting protocols and DNA barcoding and metabarcoding processing was supported by the Centre for Biodiversity Genomics, especially Michelle D’Souza and Evgeny Zakharov. All data were stored on the Earthcape Biodiversity Database Platform (https://earthcape.com/) with the support of Evgeniy Meyke. All authors were working as members of the Target Malaria Not-for-Profit Research Consortium (www.targetmalaria.org), which receives core funding from the Gates Foundation and from Coefficient Giving. DRHB is also supported by a grant from the Wellcome Trust (UNS1285360).

## Author contributions statement

BB, OTL and TDH conceived the ideas and designed methodology; BB, NAA, BA, ED and TDH collected the data with support from FAA; BB analysed the data with support from DRHB, TDH and OTL; OTL, FAA and TDH provided project supervision; BB led the writing of the manuscript. All authors contributed critically to the drafts and gave final approval for publication.

## Data archiving statement

Data is archived on the Zenodo digital repository: https://doi.org/10.5281/zenodo.19591599

