## Supplementary information for "Biting Diptera-host network structure varies with anthropogenic landscape modification"

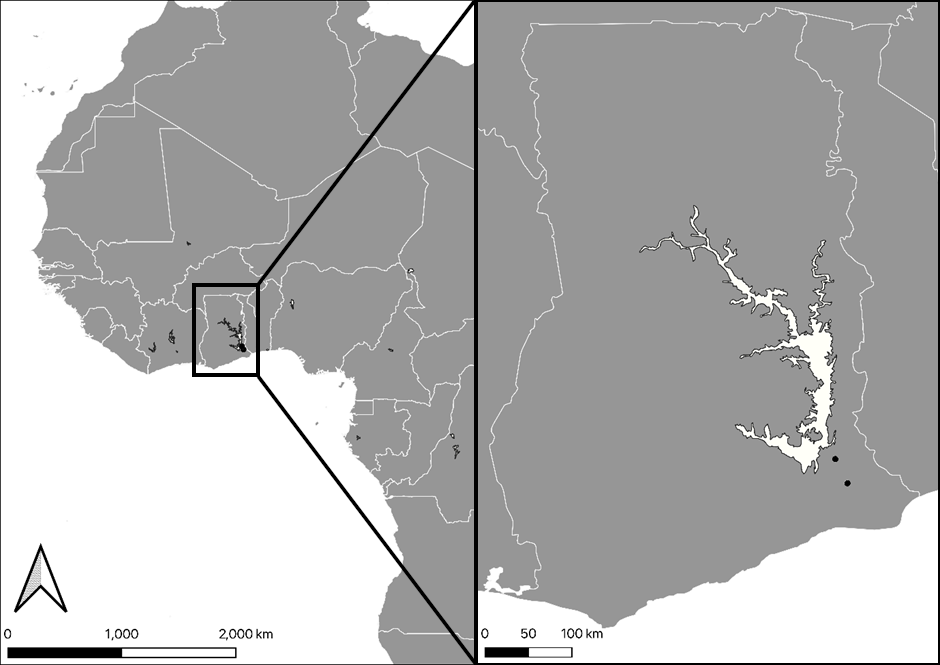

Supplementary Figure 1: Location of two study villages (black circles). Abutia Amagame (northern site) 50 km away from Mafi Agove (southern site).

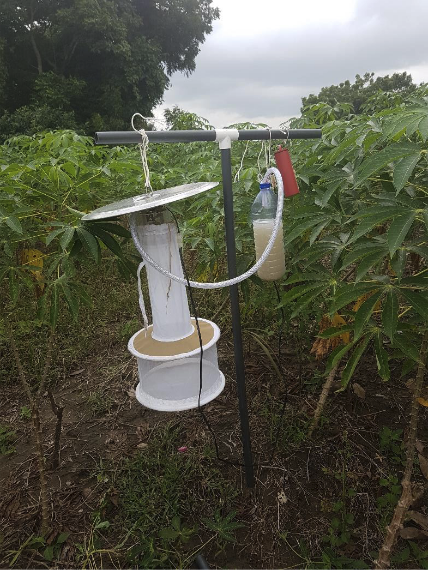

Supplementary Figure 2: The modified CDC trap with CO_2_ bait. The mesh collection area was replaced with PVC plastic containing 95% ethanol to preserve insects and blood meals

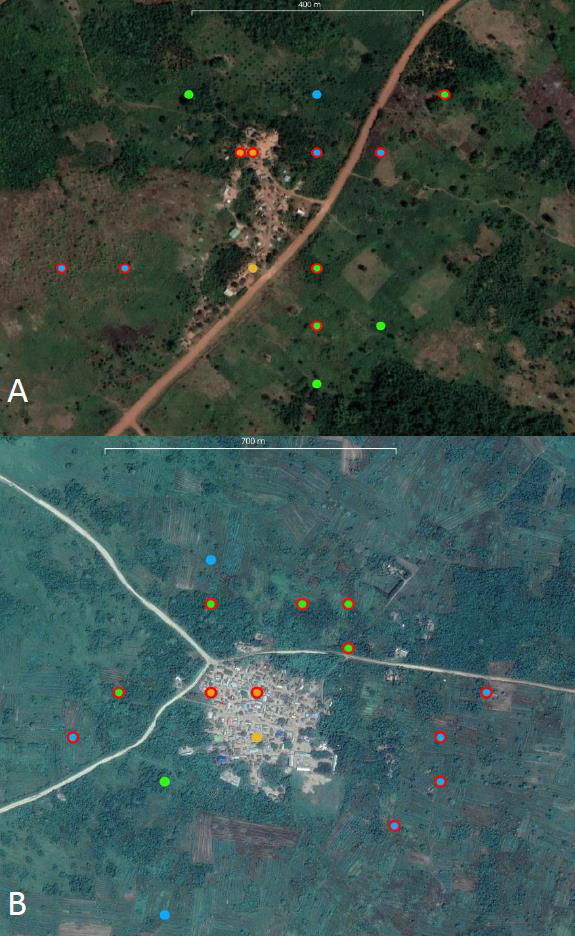

Supplementary Figure 3: Satellite image sampling locations in Abutia Amagame (A) and Mafi Agove (B) separated by habitat classification: Village (orange), Agricultural (blue), and Near-natural (green). Points bordered by a red ring indicate sites in which blood-fed Diptera were caught

Supplementary Table 1: Collection site classifications

| Lot.ID | Site | Habitat |
| --- | --- | --- |
| 955 | Abutia Amegame | Agricultural |
| 867 | Abutia Amegame | Village |
| 922 | Mafi Agove | Village |
| 779 | Abutia Amegame | Near-Natural |
| 702 | Abutia Amegame | Agricultural |
| 933 | Mafi Agove | Agricultural |
| 999 | Mafi Agove | Near-Natural |
| 1358 | Abutia Amegame | Near-Natural |
| 911 | Mafi Agove | Village |
| 1193 | Mafi Agove | Near-Natural |
| 1021 | Mafi Agove | Near-Natural |
| 1010 | Mafi Agove | Agricultural |
| 713 | Abutia Amegame | Near-Natural |
| 900 | Abutia Amegame | Near-Natural |
| 834 | Mafi Agove | Village |
| 724 | Abutia Amegame | Village |
| 1237 | Mafi Agove | Near-Natural |
| 1314 | Abutia Amegame | Village |
| 1043 | Abutia Amegame | Near-Natural |
| 1226 | Mafi Agove | Agricultural |
| 966 | Abutia Amegame | Near-Natural |
| 1109 | Mafi Agove | Near-Natural |
| 988 | Abutia Amegame | Agricultural |
| 889 | Abutia Amegame | Near-Natural |
| 1303 | Abutia Amegame | Agricultural |
| 1182 | Mafi Agove | Agricultural |
| 1032 | Mafi Agove | Agricultural |
| 878 | Abutia Amegame | Agricultural |
| 1204 | Mafi Agove | Agricultural |
| 1248 | Mafi Agove | Agricultural |

Supplementary Table 2: Non-blood-fed and blood-fed Diptera richness, number of blood meals, and host richness by location.

| Site/Habitat | Non-blood-fed Diptera richness | Non-blood-fed Diptera abundance | Blood-fed Diptera richness | Number of blood meals | Host richness |
| --- | --- | --- | --- | --- | --- |
| Abutia Amegame | 30 | 3,405 | 15 | 27 | 6 |
| Mafi Agove | 33 | 3,690 | 16 | 48 | 14 |

Supplementary Table 3: Interaction and disease competence. Each row represents a collected blood-fed Diptera species, the diseases they are competent vectors for and the susceptible hosts they interacted with, as determined by molecular blood meal analysis. Row, disease, and host order are not representative of relative importance. As we did not collect data on pathogen occurrence, these data represent hypothetical transmission events based on interactions between competent vectors and hosts.

| Species | Diseases | Hosts |
| --- | --- | --- |
| *Culex perexiguus* | Avaian malaria, West Nile virus, Rift Valley fever | *Homo sapiens* |
| *Culicoides imicola* | Bluetongue disease, African horse sickness, schmallenberg | *Homo sapiens, Bos taurus* |
| *Culicoides distinctipennis* | Bluetongue disease | *Homo sapiens, Bos taurus* |
| *Aedes bromeliae* | Chikungunya, Yellow fever | *Homo sapiens* |
| *Sergentomyia ingrami* | Leishmaniases | *Homo sapiens* |
| *Sergentomyia africana* | Leishmaniases | *Homo sapiens* |
| *Sergentomyia inermis* | Leishmaniases | *Bos taurus* |
| *Sergentomyia distincta* | Leishmaniases | *Homo sapiens* |
| *Mansonia africana* | Lymphatic filariasis, Rift Valley fever, West Nile virus | *Homo sapiens, Tyto alba* |
| *Culex decens* | Rift Valley fever, West Nile virus | *Homo sapiens* |
| *Culex univittatus* | Rift Valley fever, West Nile virus, Japanese encephalitis | *Turdus pelios* |
| *Culex cinereus* | Sindbis | *Homo sapiens* |
| *Culex watti* | West Nile virus | *Homo sapiens, Coturnix sp., Bos taurus, Pternistis bicalcaratus, Gallus gallus* |
| *Coquillettidia metallica* | West Nile virus, Middelburg, Sindbis | *Homo sapiens, Milvus migrans* |
| *Culex nebulosus* | West Nile virus, Middelburg, Ntaya | *Homo sapiens* |
| *Culex vansomereni* | West Nile virus | *Homo sapiens* |

Supplementary Table 4: Observed and estimated Diptera-host interactions and species numbers by habitat

| Site interactions and species | Observed | Estimated | S.E. | 95% Lower | 95% Upper |
| --- | --- | --- | --- | --- | --- |
| Agricultural interactions | 19 | 142.429 | 138.444 | 40.047 | 742.825 |
| Agricultural hosts | 9 | 32.625 | 30.012 | 12.46 | 170.331 |
| Agricultural Diptera | 11 | 19.679 | 9.826 | 12.463 | 62.475 |
| Near-natural interactions | 30 | 207.577 | 143.583 | 74.224 | 743.047 |
| Near-natural hosts | 11 | 22.936 | 12.829 | 13.143 | 77.49 |
| Near-natural Diptera | 22 | 84.359 | 54.579 | 36.222 | 295.42 |

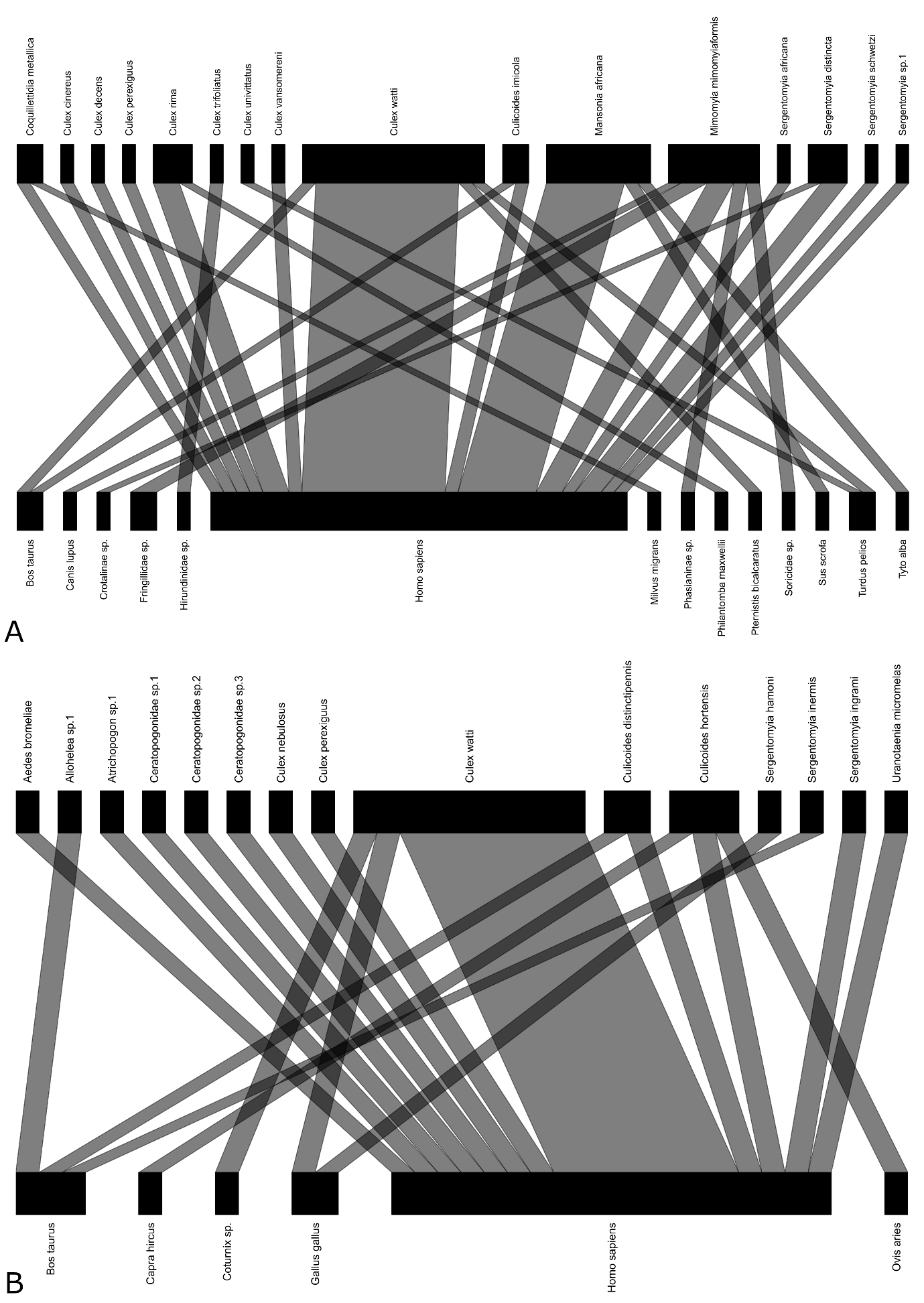

Supplementary Figure 4: Biting Diptera-host network divided by Village. Diptera (top)-host (bottom) interactions in Abutia Amagame (A) and Mafi Agove (B). Node and edge widths are proportional to frequency of occurrence.
